# Comparison of different exit scenarios from the lock-down for COVID-19 epidemic in the UK and assessing uncertainty of the predictions

**DOI:** 10.1101/2020.04.09.20059451

**Authors:** Anatoly Zhigljavsky, Roger Whitaker, Ivan Fesenko, Kobi Kremnizer, Jack Noonan

## Abstract

We model further development of the COVID-19 epidemic in the UK given the current data and assuming different scenarios of handling the epidemic. In this research, we further extend the stochastic model suggested in [1] and incorporate in it all available to us knowledge about parameters characterising the behaviour of the virus and the illness induced by it. The models we use are flexible, comprehensive, fast to run and allow us to incorporate the following:

- time-dependent strategies of handling the epidemic;
- spatial heterogeneity of the population and heterogeneity of development of epidemic in different areas;
- special characteristics of particular groups of people, especially people with specific medical pre-histories and elderly.

Standard epidemiological models such as SIR and many of its modifications are not flexible enough and hence are not precise enough in the studies that requires the use of the features above. Decision-makers get serious benefits from using better and more flexible models as they can avoid of nuanced lock-downs, better plan the exit strategy based on local population data, different stages of the epidemic in different areas, making specific recommendations to specific groups of people; all this resulting in a lesser impact on economy, improved forecasts of regional demand upon NHS allowing for intelligent resource allocation.

In this work, we investigate the sensitivity of the model to all its parameters while considering several realistic scenarios of what is likely to happen with the dynamics of the epidemic after the lock-down, which has started on March 23, is lifted.

The main findings from this research are the following

- very little gain, in terms of the projected hospital bed occupancy and expected numbers of death, of continuing the lock-down beyond April 13, provided the isolation of older and vulnerable people continues and the public carries on some level of isolation in the next 2-3 months, see Section 3.1;
- in agreement with [1], isolation of the group of vulnerable people during the next 2-3 months should be one of the main priorities, see Section 3.2;
- it is of high importance that the whole population carries on some level of isolation in the next 2-3 months, see Section 3.5;
- the timing of the current lock-down seems to be very sensible in areas like London where the epidemic has started to pick up by March 23; in such areas the second wave of epidemic is not expected, see figures in Sections 2.2 and 3;
- the epidemic should almost completely finish in July, no global second wave should be expected, except areas where the first wave is almost absent, see Section 3.4.

## 1 The model

The main differences between the model we use in this work and the model of [1] are the following:

- in the current model, we use the split into mild and severe cases of illness, see Figure 1;
- as a result, we can now better estimate the expected numbers of hospital beds required and expected deaths at time *t*; however, we still do not take into account important factors of hospital bed availability and different heterogeneities, see [1] and Sections 1.3 and 2.2;
- we include assessments of the sensitivity of our model, using Julia programming language [2]; most of the previous epidemiological models do not allow this kind of sensitivity assessment as they have far too many parameters and very rigid conditions about the probability distributions involved.

**Figure 1:**
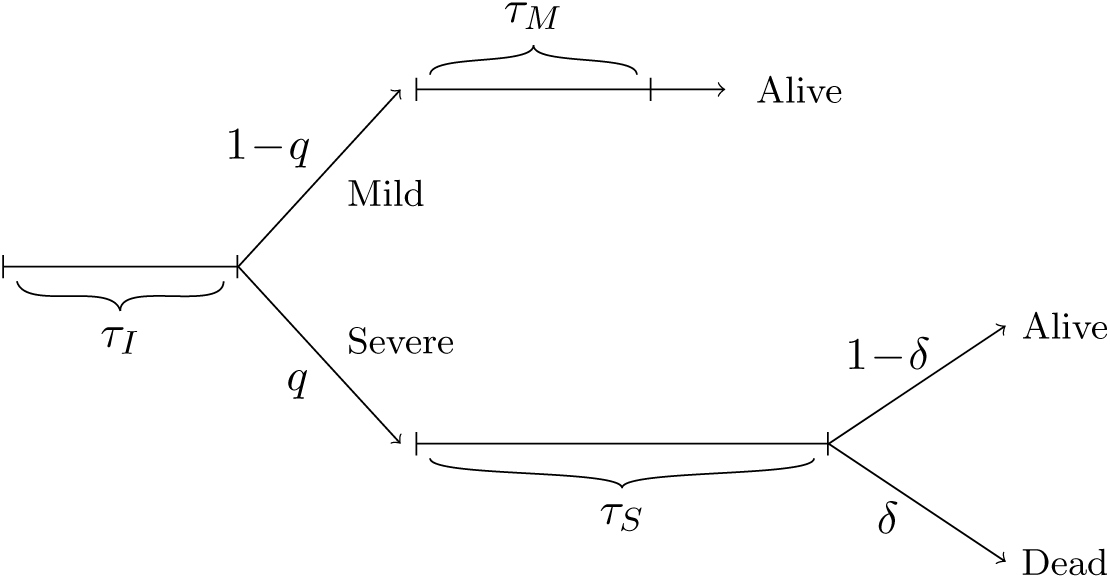
Flow-chart for the process of illness

### 1.1 Variables and parameters

- *t* - time (in days)
- *t*_1_ - the lock-down time, *t*_1_ = March 23
- *t*_2_ - time when the lock-down finishes
- *N* - population size
- *G* - sub-population (e.g. a group of people aged 70+)
- *n* - size of group *G*
- *α* = *n/N* (in the case of the UK and *G* consisting of people aged 70+, *α* = 0.132)
- *r*_*G*_ - average mortality rate in the group *G*
- *r*_*other*_ - average mortality rate for the rest of population
- *I*(*t*) - number of infectious at time *t*
- *S*(*t*) - number of susceptible at time *t*;
- 1*/σ*_*M*_ - average time for recovery in mild cases
- 1*/σ*_*S*_ - average time until recovery (or death) in severe cases
- *λ*_*I*_ - mean of the incubation period during which an infected person cannot spread the virus
- *δ* - the probability of death in severe cases
- *R*_0_ - reproductive number (average number of people who will capture the disease from one contagious person)
- *R*_1_ - reproductive number during the lock-down (*t*_1_ ≤ *t* < *t*_2_)
- *R*_2_ - reproductive number after the lock-down finishes (*t* ≥ *t*_2_)
- *c* - the degree of separation of vulnerable people
- *x* - the proportion of infected at the start of the lock-down

### 1.2 Values of parameters and generic model

The reproductive number *R*_0_ is the main parameter defining the speed of development of an epidemic. There is no true value for *R*_0_ as it varies in different parts of the UK (and the world). In particular, in rural areas one would expect a considerably lower value of *R*_0_ than in London. Authors of [3] suggest *R*_0_ = 2.2 and *R*_0_ = 2.4 as typical; the authors of [4] use values for *R*_0_ in the range [2.25, 2.75]. We shall use the value *R*_0_ = 2.5 as typical, which, in view of the recent data, looks to be a rather high (pessimistic) choice overall. However, *R*_0_ = 2.5 seems to be an adequate choice for the mega-cities where the epidemics develop faster and may lead to more causalities. In rural areas, in small towns, and everywhere else where social contacts are less intense, the epidemic is milder.

The flow-chart in Figure 1 describes the process of illness. We assume that the person becomes infected *τ*_*I*_ days after catching the virus, where *τ*_*I*_ has Poisson distribution with mean of *λ*_*I*_ days. Parameter *λ*_*I*_ defines the mean of the incubation period during which the person cannot spread the virus. Recommended value for *λ*_*I*_ is *λ*_*I*_ = 5.

We then assign probabilities *q* and 1 − *q* for a person to get a severe or mild case correspondingly. The value of *q* depends on whether the person belongs to the group *G* (then *q* = *q*_*G*_) or the rest of population (in this case, *q* = *q*_*other*_). In Section 1.3 below we will relate the probabilities *q*_*G*_ and *q*_*other*_ to the mortality rates in the two groups.

In a mild case, the person stays infectious for *τ*_*M*_ days and then discharges alive (that is, stops being infectious). We assume that the continuous version of *τ*_*M*_ has Erlang distribution with shape parameter *k*_*M*_ and rate parameter *λ*_*M*_, the mean of this distribution is *k*/*λ*_*M*_ = 1*/σ*_*M*_ (in simulations, we discretise the numbers to their nearest integers). We use values *k*_*M*_ = 3 and *λ*_*M*_ = 1*/*2 so that *Eτ*_*M*_ = *k*_*M*_ */λ*_*M*_ = 1*/σ*_*M*_ = 6. The variance of *τ*_*M*_ is 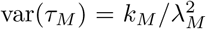 for *k*_*M*_ = 3 and *λ*_*M*_ = 1*/*2 we have var(*τ*_*M*_) = 12 which seems to be a reasonable value.

In a severe case, the person stays infectious for *τ*_*S*_ days. The continuous version of *τ*_*S*_ has Erlang distribution with shape parameter *k*_*S*_ and rate parameter *λ*_*S*_. The mean of this distribution is *k*_*S*_*/λ*_*S*_ = 1*/σ*_*S*_. We use values *k*_*S*_ = 3 and *λ*_*S*_ = 1*/*7 so that *Eτ*_*S*_ = *k*_*S*_*/λ*_*S*_ = 21, in line with the current knowledge, see e.g. [5, 6, 7]. The variance of *τ*_*S*_ is 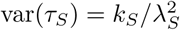 for *k*_*M*_ = 3 and *λ*_*S*_ = 1*/*7 the standard deviation of the chosen Erlang distribution is approximately 12, which is rather large and reflects the uncertainty we currently have about the period of time a person needs to recover (or die) from COVID-19.

In a severe case, the person dies with probability *δ* > 0 on discharge. The relation between *δ*, the average mortality rates, the probabilities *q*_*S*_ for the group *G* and the rest of population is discussed in Section 1.3.

Note that the use of Erlang distribution is standard for modelling similar events in reliability and queuing theories, which have much in common with epidemiology.

### 1.3 Probability of getting a severe case, probability of death in case of severe case *δ* and mortality rates

In this section, we relate the probability of getting severe case from group *G* and the rest of population to the average mortality rates.

We define *G* as the group of vulnerable people. In the computations below, we assume that *G* consists of people aged 70+. We would like to emphasise, however, that there is still a lot of uncertainty on who is vulnerable. Moreover, it is very possible that the long term effect of the virus might cause many extra morbidities in the coming years coming from severe cases who recover.

We use the common split of the UK population into following age groups:

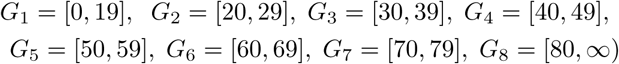

and corresponding numbers *N*_*m*_ (*m* = 1, …, 8; in millions) taken from [8]

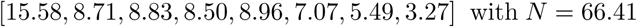

The probabilities of death for group *G*_*m*_, denoted by *p*_*m*_, are given from Table 1 in [3] and replicated many times by the BBC and other news agencies are:

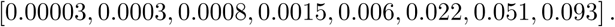

Unfortunately, these numbers do not match the other key number given in [3]: the UK average mortality rate which is estimated to be about 0.9%. As we consider the value of the UK average mortality rate as more important, we have multiplied all probabilities above by 0.732 to get the average mortality rate to be 0.9%.

Moreover, recent COVID-19 mortality data shows that the average mortality rates, especially for younger age groups, are significantly lower than the ones given above. As we do not have reliable sources for the average mortality rates, we shall use the data from [3] adjusted to the average mortality rate to be 0.9%. As we shall be getting better estimates of the average mortality rates, the model can be easily adjusted to them.

There is an extra controversy which concerns the definition of what is death caused by COVID-19. As an example, the following is a direct quote from the official ONS document [9]: *“In Week 13, 18*.*8% of all deaths mentioned “Influenza or Pneumonia”, COVID-19, or both. In comparison, for the five-year average, 19*.*6% of deaths mentioned “Influenza and Pneumonia”. “Influenza and Pneumonia” has been included for comparison, as a well-understood cause of death involving respiratory infection that is likely to have somewhat similar risk factors to COVID-19*.*”* In view of facts like this, the true COVID-19 mortality rates could be up to 5-10 times lower than given in [3] and used in this work.

If consider population data for the whole UK and define the group *G* as a union of groups *G*_7_ and *G*_8_, we have *n* = 5.49 + 3.27 = 8.76m and *α* = 8.76*/*66.41 ≃ 0.132. The mortality rate in the group *G* and for the rest of population are therefore

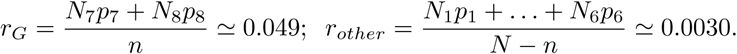

Consider now a random person from the group *G* who has got an infection. The probability he dies is *r*_*G*_. On the other hand, this is also the product of the probability that this person has a severe case (which is *q*_*G*_) and the probability (which is *δ*) that the person dies conditionally he/she has severe case. The same is true for the rest of population. Therefore,

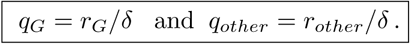

The COVID-19 epidemic in the whole of the UK is subject to different heterogeneities discussed in detail in [1]. The model above assumes homogeneity and hence cannot be applied to the whole of the UK and we feel it would be irresponsible to estimate the total death toll for the UK using such a model. Instead, we shall assume that we apply this model to the population of inner London with population size rounded to 3 million. It may be tempting to multiply our expected death tolls by 22=66/3 but this would give very elevated forecasts. Indeed, epidemic in inner London can be considered as the worst-case scenario and, in view of [1], we would recommend to down-estimate the UK overall death toll forecasts by a factor of 2 or even more.

## 2 Main scenarios

In this section we formulate two main scenarios and explain the main figures. In Section 3 we study sensitivity with respect to all model parameters.

### 2.1 Two plots for the main set of parameters, no intervention

As the main set of parameters for the main model we use the following:

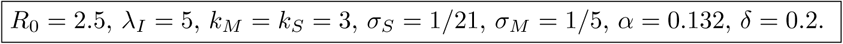

Note that in Section 2.2 we shall introduce four further parameters which will describe the lock-down of March 23 and the situation at the exit from this lock-down.

In Figure 2 we plot, in the case with no intervention, proportions of infected at time *t* (either overall or in the corresponding group) and use the following colour scheme:

**Figure 2:**
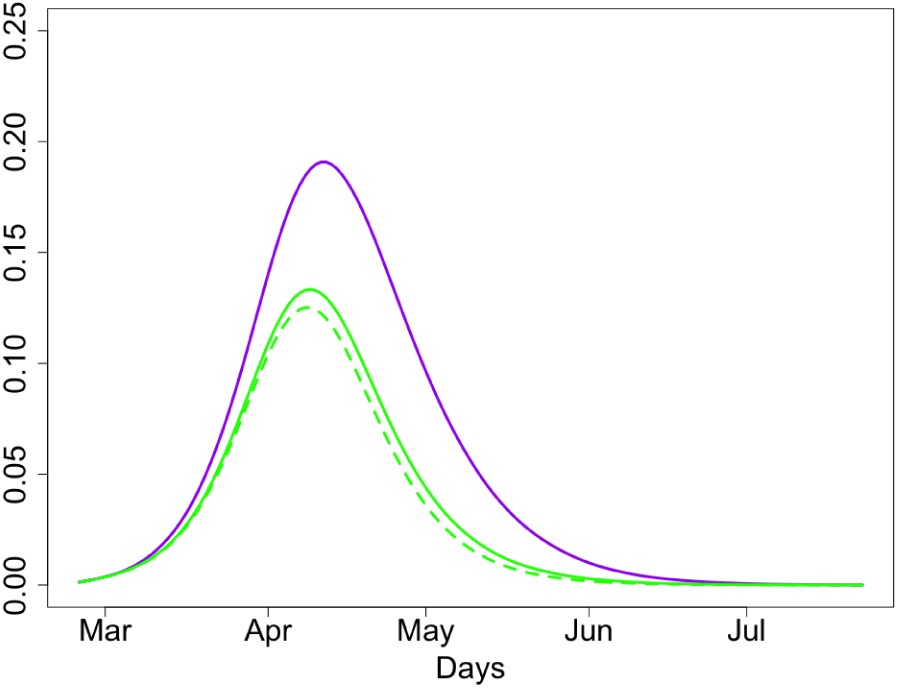
Proportions of people infected at time *t* (overall, group *G* and the rest of population).

- Solid green: *I*(*t*), the proportion of infected in overall population at time *t*.
- Purple: *I*_*G*_(*t*), the proportion of infected in group *G* at time *t*
- Dashed green: the proportion of infected in the rest of population.

Knowing proportion of infected at time *t* is important for knowing the danger of being infected for non-infected and non-immune people.

In Figure 3 we plot, in the case of no intervention, proportions of people with severe case of disease at time *t* and proportions of such people discharged at time *t*; there are separate plots for the group *G* and the rest of population.

**Figure 3:**
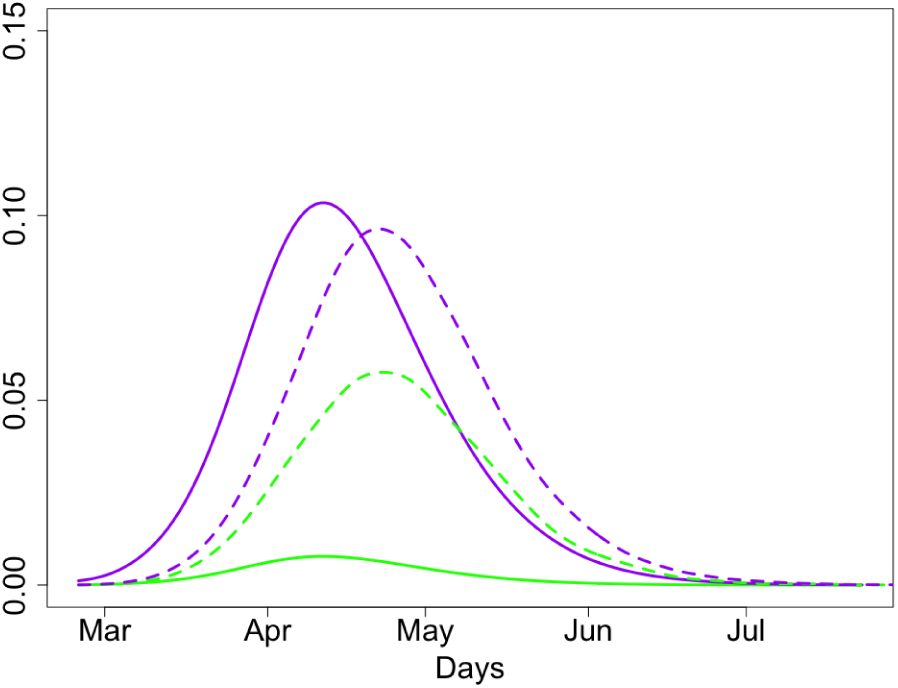
Proportions of people with severe case of disease at time *t* (solid lines) and died at time *t* (dashed lines).

- Solid purple: proportion of people from group *G* with severe case at time *t*.
- Solid green: proportion of people from the rest of population with severe case at time *t*.
- Dashed purple: proportion of people from group *G* died at time *t*, multiplied by 100.
- Dashed green: proportion of people from the rest of population died at time *t*, multiplied by 1000 (so that the curve can be seen in the figure).

Proportions of people with severe case at time *t* is the main characteristic needed for planning NHS work-load. The proportion of people with severe case from group *G* discharged at time *t* is proportional to the expected number of deaths after multiplying this number by *δ* and the size of the group *G*; similar calculations can be done for the rest of population. Note that in these calculations we do not take into consideration an extremely important factor of hospital bed availability are at time *t*. Currently, we do not have a model for this.

We provide the values of the overall expected death toll. In the case of no lock-down, this toll would be 24(7+17)K, where the first number in the bracket correspond to the rest of population and the second one to the group *G*.

### 2.2 Key plots for the main set of parameters, the case with intervention

In this section, we study the scenario where we have made an intervention on *t*_1_ = March 23 by reducing *R*_0_ to *R*_1_ < *R*_0_ and then terminating the lock-down on *t*_2_ = April 22.

As we do not know at which stage of the epidemic the lock-down has started, we set a parameter to denote it. This parameter is a number *x* ∈ (0, 1) such that *S*(*t*_1_)*/N* = *x*.

After lifting the lock-down, we assume that people from the group *G* are still isolated but the rest of population returns back to normal life. However, it is natural to assume that the reproduction number *R*_2_ for *t* ≥ *t*_2_ is smaller that the initial value of *R*_0_. To denote the degree of isolation of people from *G* for *t* ≥ *t*_2_, we assume that for *t* ≥ *t*_2_ the virus is transmitted to people in such a way that, conditionally a virus is transmitted, the probability that it reaches a person from *G* is *p* = *cα* with 0 < *c* ≤ 1. Our main value for *c* is 1*/*4. This means that for *t* ≥ *t*_2_, under the condition that a virus is infecting a new person, the probabilities that this new person belongs to *G* is 1*/*5.

Measuring the level of compliance in the population and converting this to simple epidemiological measures *c, R*_1_ and *R*_2_ and is hugely complex problem which is beyond the scope of this paper.

Summarizing, there are five parameters modelling the lock-down:

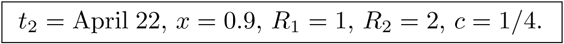

Figures 4 and 5 similar to Figures 2 and 3 but computed for the case of intervention. The colour schemes in Figures 4 and 5 are exactly the same as in Figures 2 and 3, respectively. The two grey lines in Figures 4 and 5 mark the intervention times. Note different scales in the *y*-axis in Figures 4 and 5 in comparison to Figures 2 and 3

**Figure 4:**
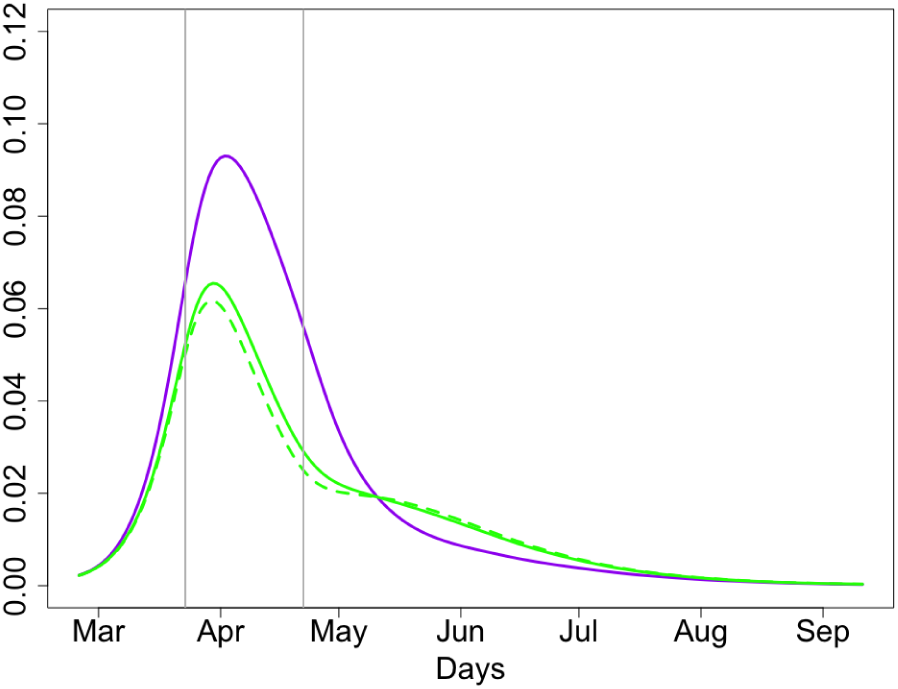
Proportions of people infected at time *t* (overall, group *G* and the rest of population).

**Figure 5:**
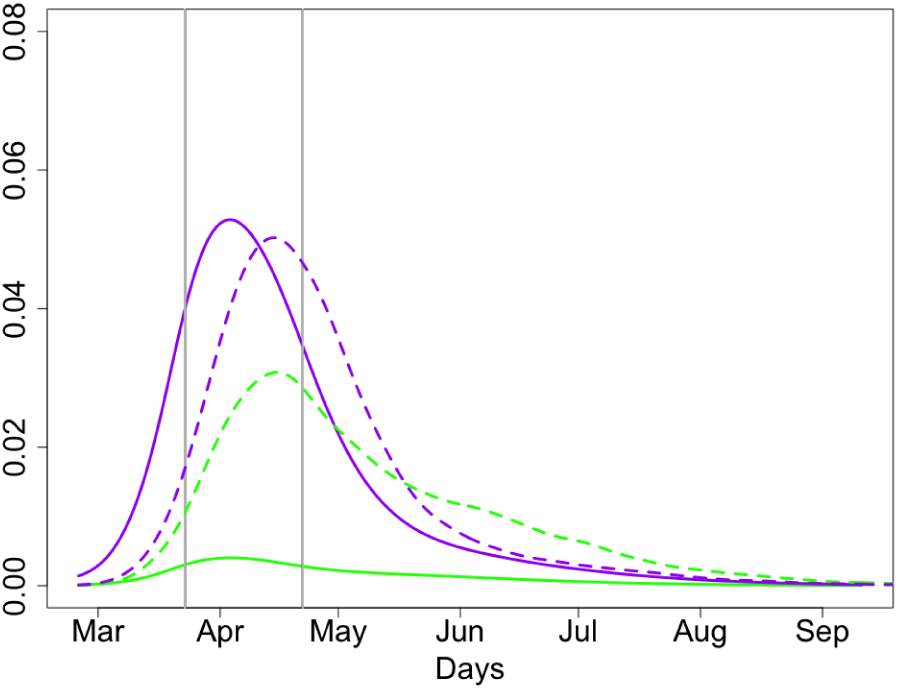
Proportions of people with severe case of disease at time *t* (solid lines) and discharged at time *t* (dashed). Expected death toll: 15K

We will study sensitivity of the curves *I*(*t*) and *I*_*G*_(*t*), plotted in Figures 4 and again in Figure 6 for the same parameters, with respect to all parameters of the model. In Figure 7 we plot average death numbers (purple for group *G* and green for the rest of the population) in the case of the inner London as explained at the end of Section 1.3. Note that these numbers do not take into account the important factor of hospital bed availability and various heterogeneities, as explained in [1].

**Figure 6:**
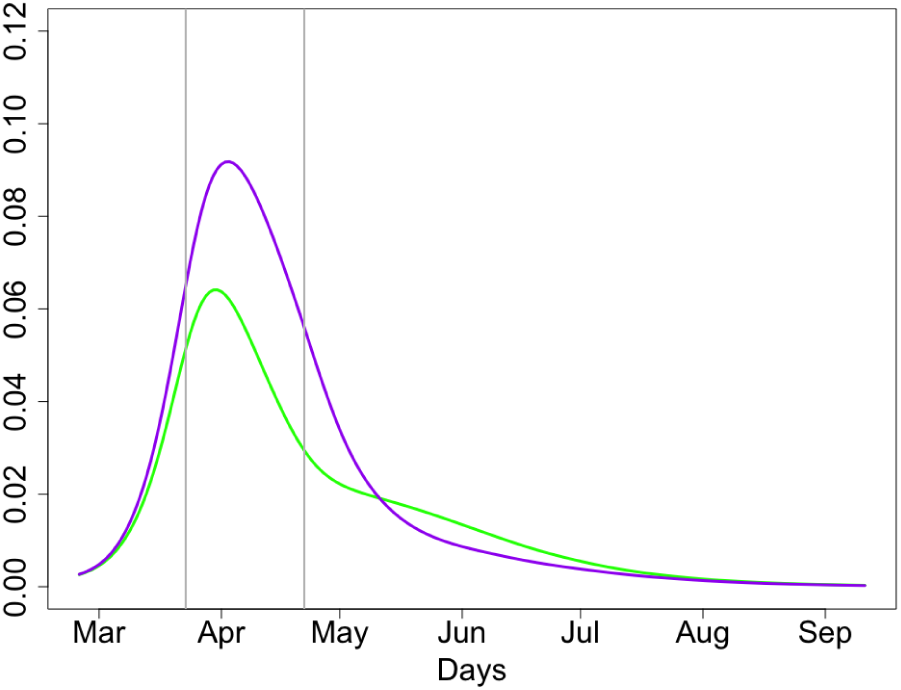
Proportions of people infected at time *t* (group *G* and overall).

**Figure 7:**
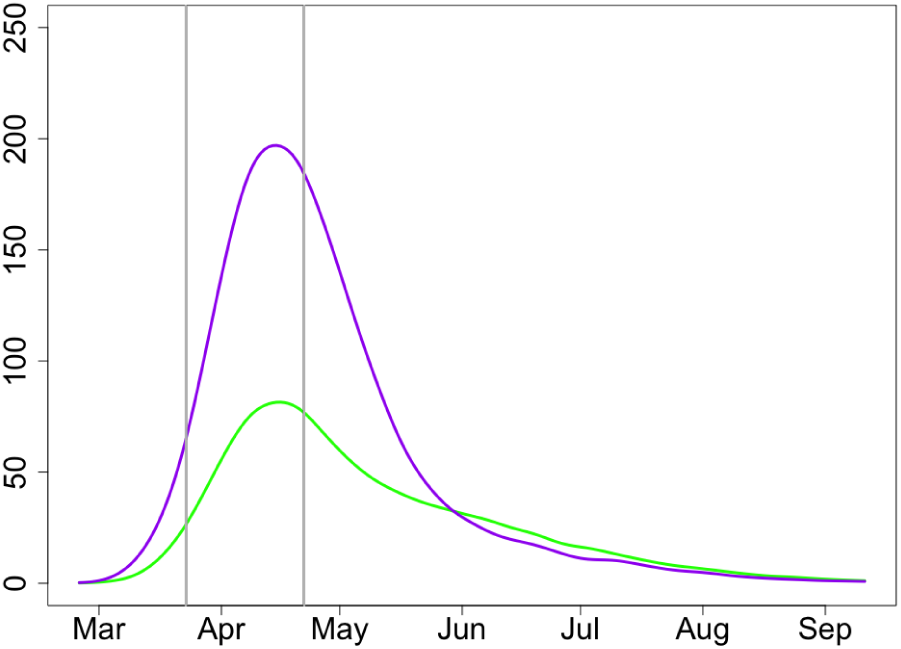
Expected deaths at time *t* at group *G* and the rest of population.

## 3 Sensitivity to main parameters of the model and exit strategy

We have selected Figures 6 and 7 as the main figures. The next pairs of figures show sensitivity of the model with respect to different parameters in the model. All these pairs of figures are similar Figures 6 and 7 except one of the parameters is changing. Moreover, all the curves from Figures 6 and 7 are reproduced in the pairs of figures below in the same colours.

The colour scheme in all figures below with even numbers is:

- Solid purple: *I*_*G*_(*t*)*/n*, the proportion of infected at time *t* from group *G*; main set of parameters;
- Solid green: *I*(*t*)*/N*, the proportion of infected at time *t* from the population; main set of parameters;
- Solid (and perhaps, dashed) red: *I*_*G*_(*t*)*/n* for the alternative value (values) of the chosen parameter;
- Solid (and perhaps, dashed) blue: *I*(*t*)*/N* for the alternative value (values) of the chosen parameter. The colour scheme in all figures below with odd numbers is:
- Solid purple: *D*_*G*_(*t*), the expected number of death at time *t* at group *G*; main set of parameters;
- Solid green: *D*_*other*_(*t*), the expected number of death at time *t* for the rest of population; main set of parameters;
- Solid (and perhaps, dashed) red: *D*_*G*_(*t*) for the alternative value (values) of the chosen parameter;
- Solid (and perhaps, dashed) blue: *D*_*other*_(*t*) for the alternative value (values) of the chosen parameter.

### 3.1 Sensitivity to *t*_2_, the time of lifting the lock-down

In Figures 8 and 9, we have moved the time *t*_2_, the time of lifting the lock-down restrictions, 9 days forward so that the lock-down period is 21 days rather than 30 days as in the main scenario. The solid red and blue colours in Figures 8 and 9 correspond to *t*_2_ = April 13 and the middle grey vertical line marks April 13.

**Figure 8:**
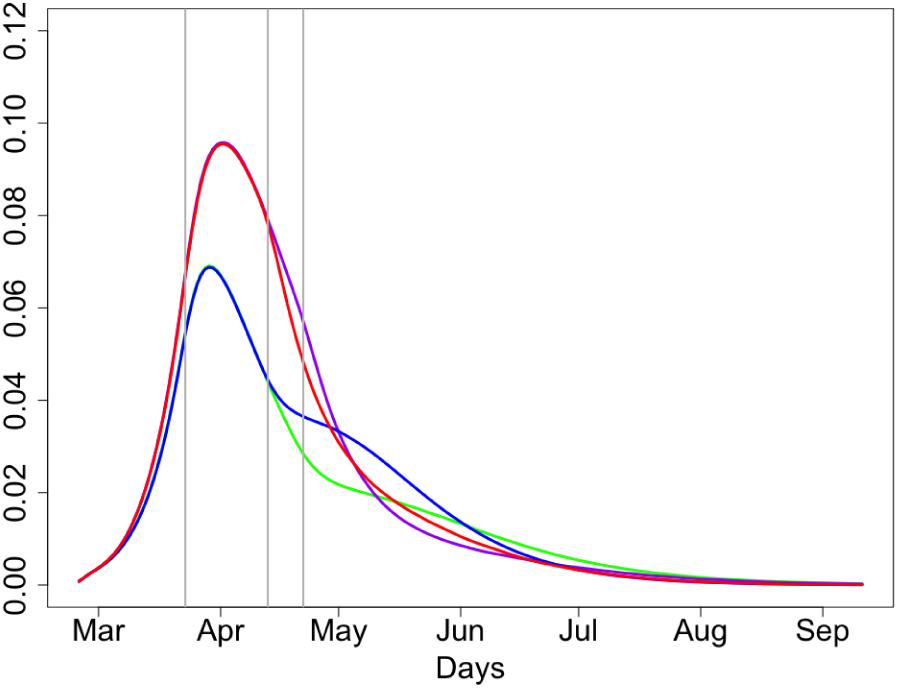
Proportions of people infected at time *t*; *t*_2_ = April 13, 22

**Figure 9:**
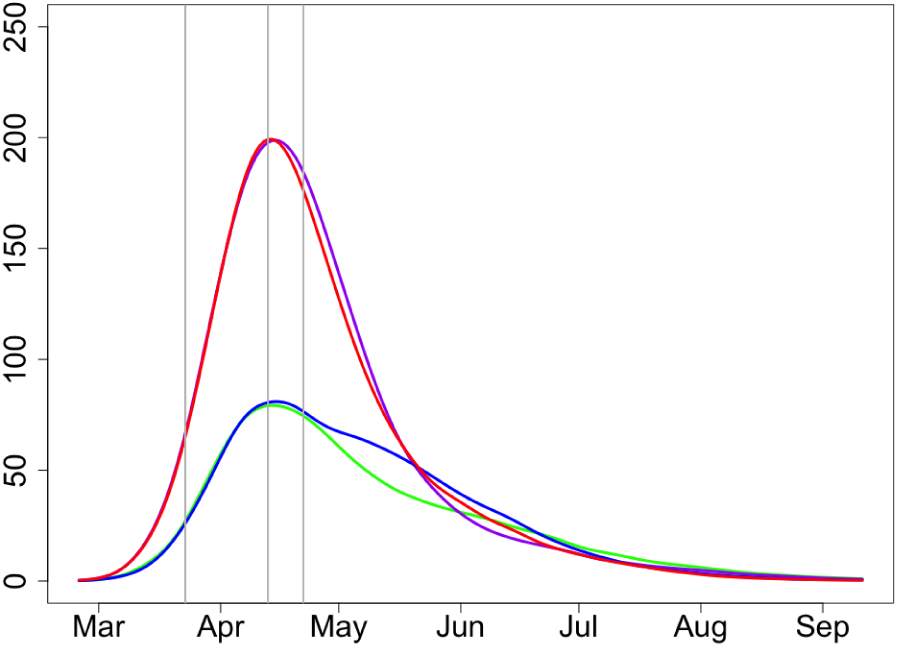
Expected deaths at time *t* at group *G* and the rest of population; *t*_2_ = April 13, 22

Consider first Figure 8 showing proportions of infected people and reflecting the demand for hospital beds. Since in the scenario *t*_2_ = April 13 we start isolating people from *G* earlier than for *t*_2_ = April 22, the number of infected from group *G* (red solid line) is slightly lower during the end of April – beginning of May for *t*_2_ = April 13 (purple solid line). The total number of infections from the rest of population is clearly slightly higher for *t*_2_ = April 13 (blue line) than for *t*_2_ = April 22. Taking into account the fact that infected people from *G* would on average require more hospital beds than the rest of population, the expected number of hospital beds required for the whole population stays approximately the same during the whole epidemic which has to be almost over in July.

Figure 9, showing expected deaths numbers for both scenarios with *t*_2_ = April 13 and *t*_2_ = April 22, show similar patterns concluding that there is very little gain in keeping the lock-down, especially taking into account huge economic loss caused by every extra day of the lock-down [10]. Overall, the total expected death is higher but the difference can be considered as very small. Expected deaths tolls for *t*_2_ = April 22 and *t*_2_ = April 13 are 15(5.2+9.8)K and 15.3(5.6+9.7)K, respectively.

#### Conclusion

*The lock-down started on March 23 has slowed the speed of COVID-19 epidemic. However, there is very little gain, in terms of the projected hospital bed occupancy and expected numbers of death, of continuing the lock-down beyond April* 13.

### 3.2 Sensitivity to *c*, the degree of separation of people from group *G*

In Figures 10 and 11, solid and dashed line styles (for red/blue colours) correspond to *c* = 1 and *c* = 0.5 respectively. By increasing *c* we significantly increase the death toll in the group *G*. It is clear that the value of *c* measures the degree of isolation of people from *G* and has negligible effect on the rest of population; this is clearly seen in both figures. On the other hand, the value of *c* has very significant effect on the people from *G*.

**Figure 10:**
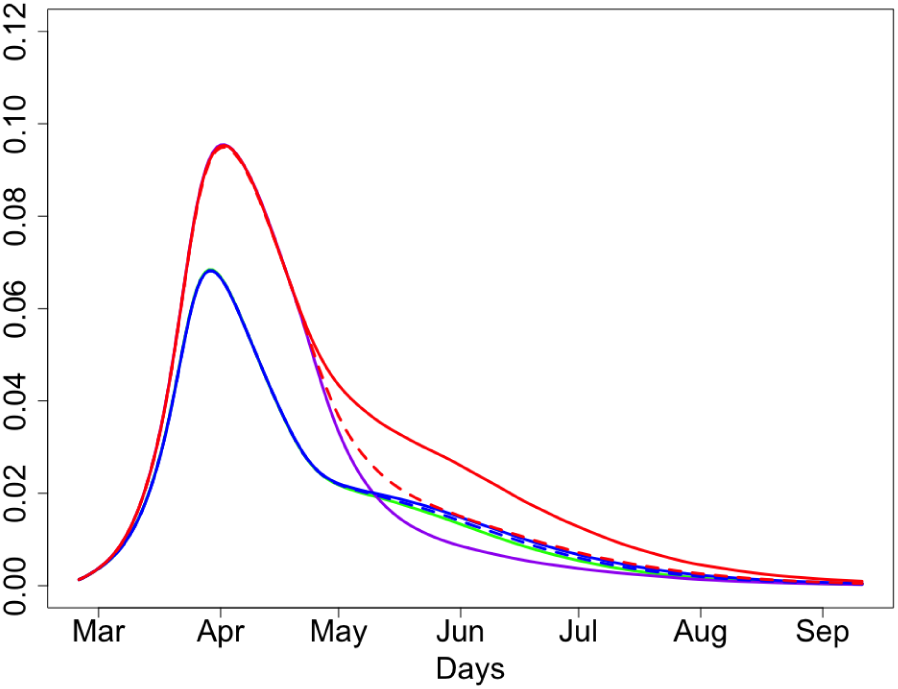
Proportions of people infected at time *t*; *c* = 0.25, 0.5, 1

**Figure 11:**
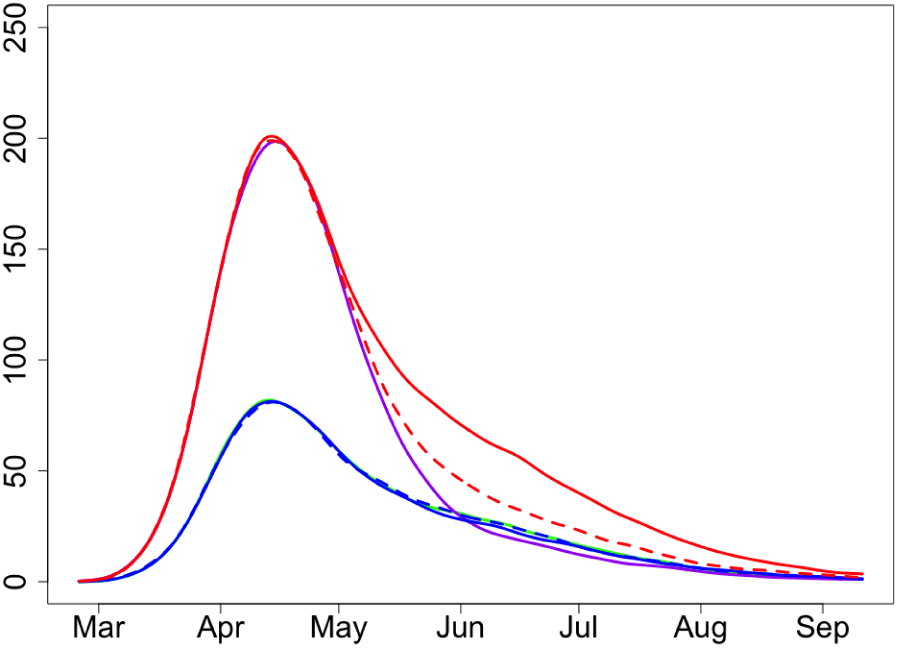
Expected deaths at time *t* at group *G* and the rest of population; *c* = 0.25, 0.5, 1

Consider first Figure 10 giving proportions of infected people. One can clearly see that the purple line (group *G, c* = 0.25) is much lower that the solid red line (group *G, c* = 1) for all the time until the end of the epidemic, from May until August. In June, in particular, the number of infected people from *G* with *c* = 0.25 is about 40% of the number of infected people from *G* with *c* = 1.

Curves on Figure 11, showing expected deaths at time *t* in group *G* and the rest of population, follow the same patterns (with about 2-week time shift) as the related curves of Figure 10 displaying the number of infected. In June and July we should expect only about 40% of the number of deaths in the scenario with isolation of people from *G* (*c* = 0.25) relative to the scenario with no special isolation for people from *G* (*c* = 1).

This is reflected in the expected deaths tolls for entire period of epidemic. Indeed, expected deaths tolls for *c* = 1 and 0.5 are 18.2(5.2+13)K and 16.2(5.2+11)K, respectively; compare this with 15(5.2+9.8)K for *c* = 0.25.

#### Conclusion

*The effect of the degree of isolation of people from group G in the aftermath of the lock-down is very significant*.

### 3.3 Sensitivity to *R*_0_

In view of recent data, the value of *R*_0_ is likely to be lower than 2.5, especially in small towns and rural areas. As Figures 12 and 13 demonstrate, in such places the epidemic will be significantly milder. The overall dynamics of the epidemic would not change much though.

**Figure 12:**
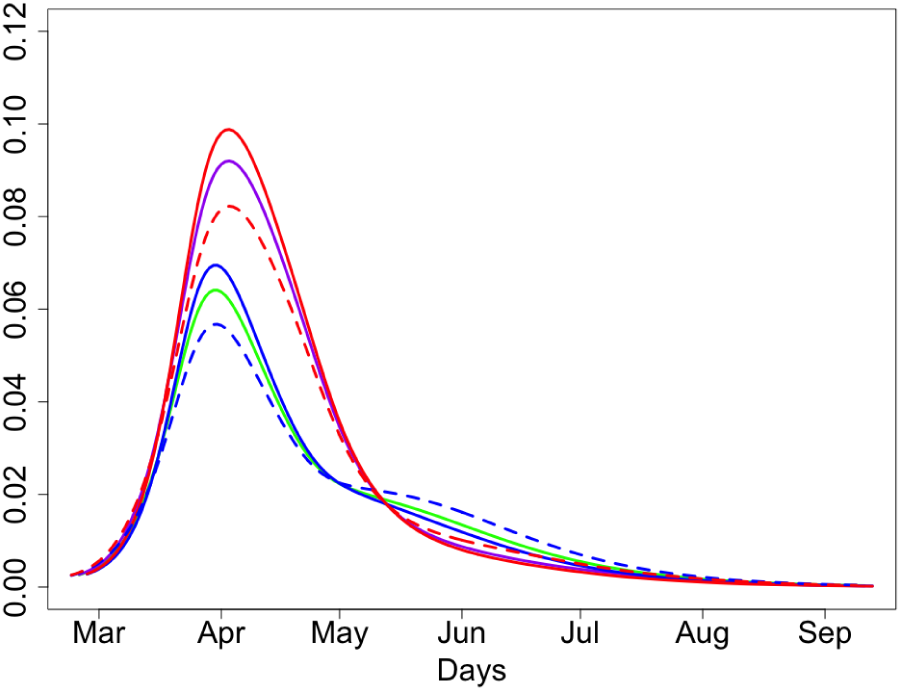
Proportions of people infected at time *t*; *R*_0_ = 2.3, 2.5, 2.7

**Figure 13:**
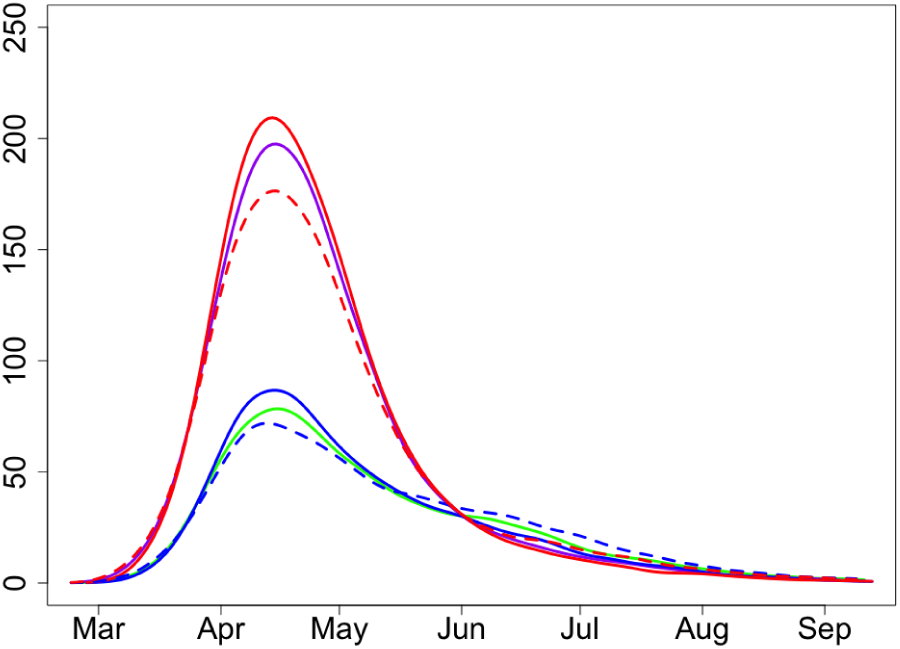
Expected deaths at time *t* at group *G* and the rest of population; *R*_0_ = 2.3, 2.5, 2.7

In Figures 12 and 13, solid and dashed line styles (for blue/red colours) correspond to *R*_0_ = 2.3 and *R*_0_ = 2.7 respectively. Expected deaths tolls for *R*_0_ = 2.3, 2.5 and *R*_0_ = 2.7 are 14.5(5+9.5)K, 15(5.2+9.8)K and 15.5(5.3+10.2)K, respectively.

### 3.4 Sensitivity to *x*, the proportion of infected at the start of the lock-down

In Figures 14 and 15, we use *x* = 0.8 and *x* = 0.9. Expected deaths toll for *x* = 0.8 is 17.3(5.3+12)K. This is higher than 15(5.2+9.8)K for *x* = 0.9. The fact that the difference is significant is related to the larger number of death for *x* = 0.8 in the initial period of the epidemic. In Figure 14, red and blue display the number of infected at time *t* for *x* = 0.8 for people from group *G* and the rest of population, respectively. Similarly, in Figure 15 the red and blue colours show the expected numbers of deaths in these two groups. Figures 14 and 15 show that the timing of the lock-down has serious effect on the development of the epidemic. Late call for a lock-down (when *x* = 0.8) helps to slow down the epidemic and guarantees its fast smooth decrease but does not safe as many people as, for example, the call with *x* = 0.9.

**Figure 14:**
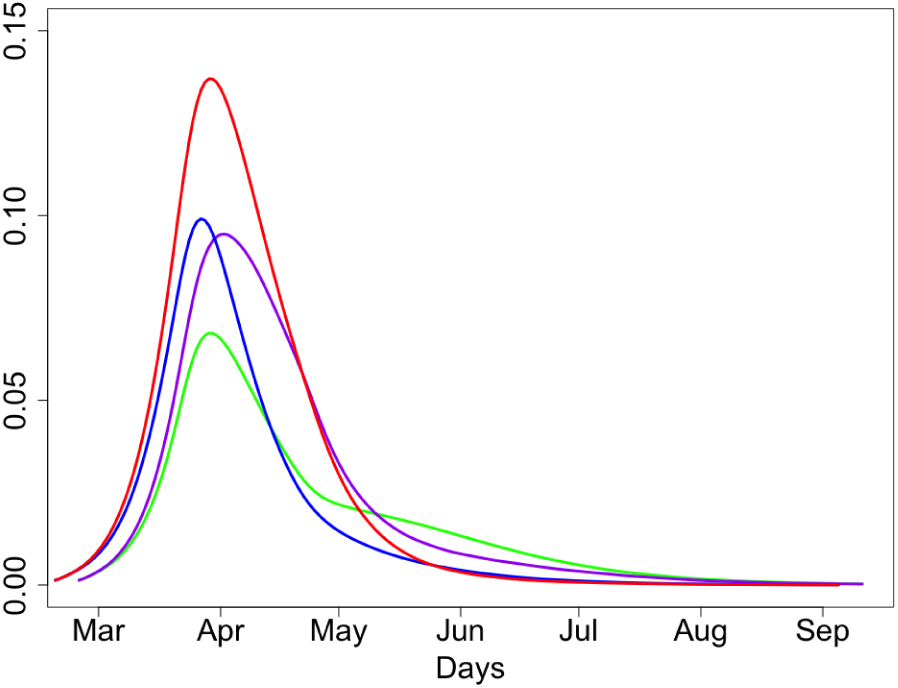
Proportions of people infected at time *t*; *x* = 0.8, 0.9

**Figure 15:**
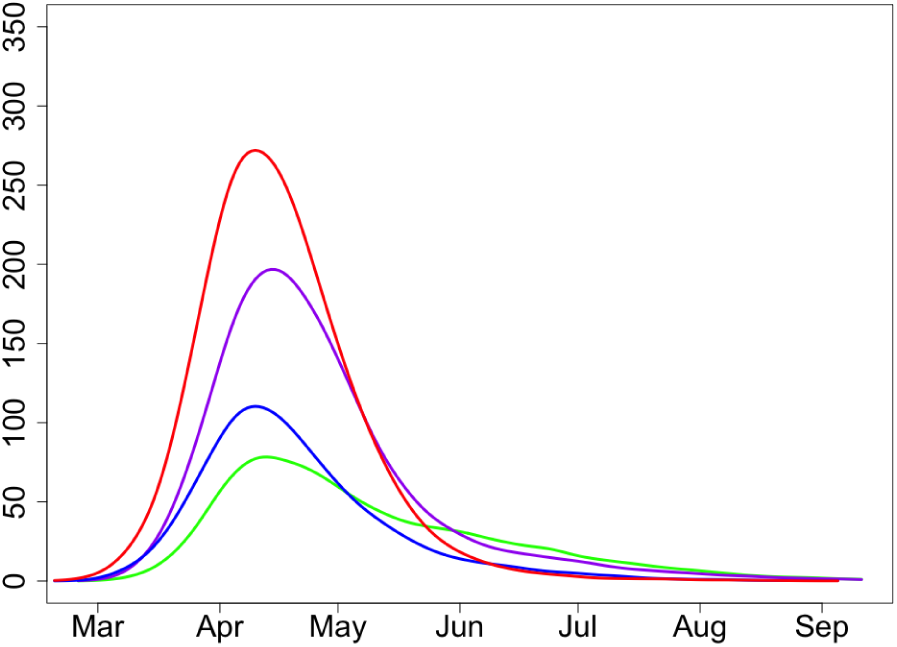
Expected deaths at time *t* at group *G* and the rest of population; *x* = 0.8, 0.9

Note that in both cases, when *x* = 0.8 and *x* = 0.9, the second wave of the epidemic is not expected as by the time of lifting the lock-down, a large percentage of the population (about 25% in case *x* = 0.9 and almost 40% in case *x* = 0.8) is either infected or immune and the ‘herd immunity’ would follow shortly. In a certain sense, the decision of making a lock-down at around *x* = 0.9 seem to be a very sensible decision to make (considering economic costs of each day of a lock-down) as this saves many lives and guarantees smooth dynamics of the epidemic with no second wave.

Let us now consider more informative scenarios when *x* = 0.95 and *x* = 0.97; that is, when the lock-down is made early, see Figures 16,17 for *x* = 0.95, 0.9 and Figures 18,19 for *x* = 0.97, 0.9. With an early lock-down, there is a clear gain in hospital bed occupancy and expected number of death at the first stage of the epidemic. However, the second wave of the epidemic should be expected in both cases *x* = 0.95 and *x* = 0.97 with a peak at around 2 months after the first one, and the second peak could be higher than the first one. This can be explained by observing that, even after 2 months of an epidemic with large *x*, even with lock-down and strong isolation of the group *G* and relatively small reproductive number (recall *R*_2_ = 2), there is still a very large proportion of non-immune people available for the virus; a large part of these people is going to be infected even with smaller reproductive number. This prolongs the epidemics. Expected deaths toll for *x* = 0.95 is 13.8(5.5+8.3)K and for *x* = 0.97 it is 13.4(5.7+7.7)K. These numbers are naturally lower than 15(5.2+9.8)K for *x* = 0.9 but the difference is not significant.

**Figure 16:**
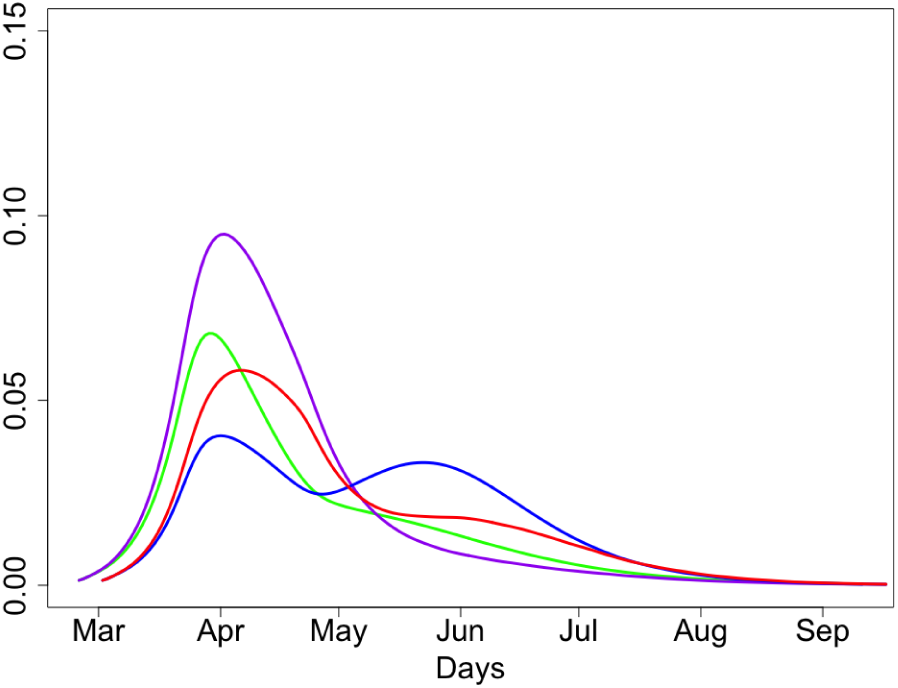
Proportions of people infected at time *t*; *x* = 0.9, 0.95

**Figure 17:**
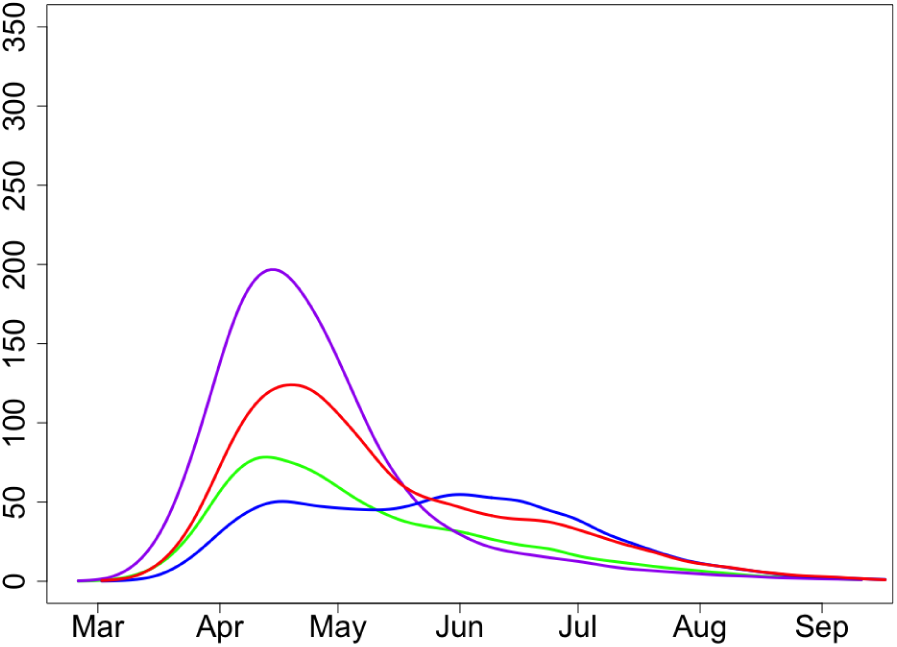
Expected deaths at time *t* at group *G* and the rest of population; *x* = 0.9, 0.95

**Figure 18:**
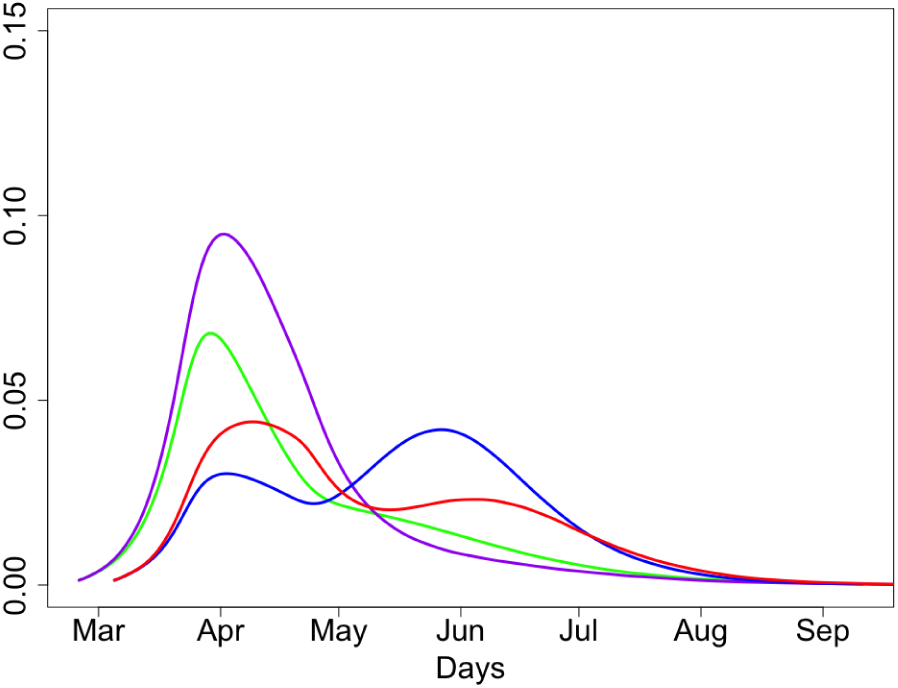
Proportions of people infected at time *t*; *x* = 0.9, 0.97

**Figure 19:**
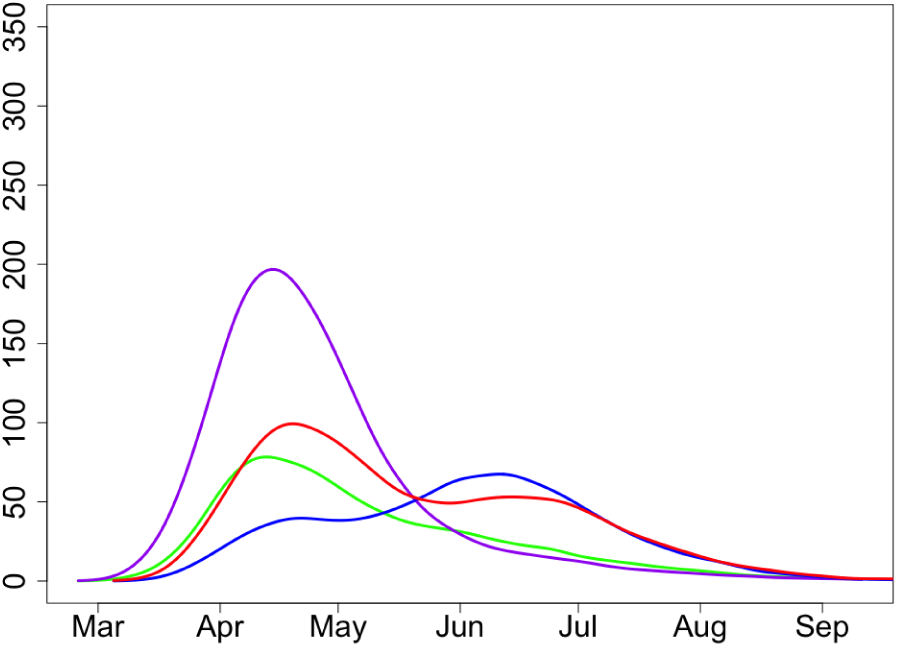
Expected deaths at time *t* at group *G* and the rest of population; *x* = 0.9, 0.97

Figures 16–19 and the discussion above imply the following conclusion.

#### Conclusion

*A lock-down at an early stage of an epidemic* (*unless it is a very strict one like in Wuhan*) *is not a sensible decision in view of the economic consequences of the lock-down and the measures required for all the remaining* (*much longer*) *period of the epidemic. Moreover, in the case of an early lock-down, the second wave of the epidemic should be expected with a peak at around 2 months after the first one*.

In Figures 18 and 19, we use *x* = 0.97 and *x* = 0.9. Expected deaths toll for *x* = 0.97 is 13.8(5.7+8.3)K. This is naturally lower than 15(5.2+9.8)K for *x* = 0.9. This is related to the fact that we isolate people from group *G* much earlier.

A disadvantage of this scenario is the second wave of epidemic with a peak at around 2 months after the first one.

### 3.5 Sensitivity to *R*_2_, the reproductive number after lifting the lock-down

In Figures 20 and 21, we use *R*_2_ = 2.0 and 2.5. Increase in *R*_2_ would lead to a significant increase in the number of severe cases and expected death numbers.

**Figure 20:**
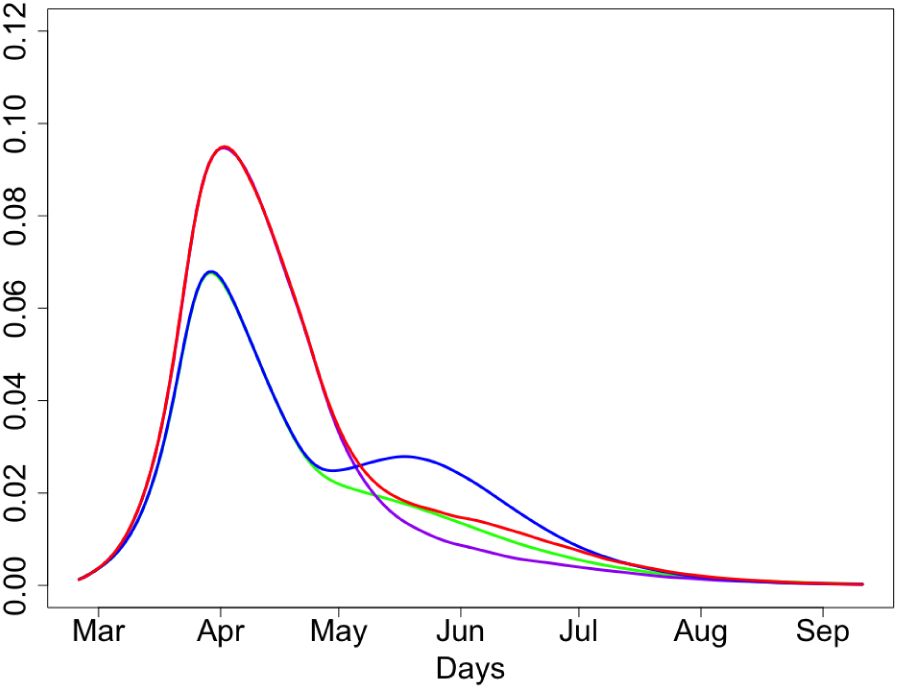
Proportions of people infected at time *t*; *R*_2_ = 2, 2.5

**Figure 21:**
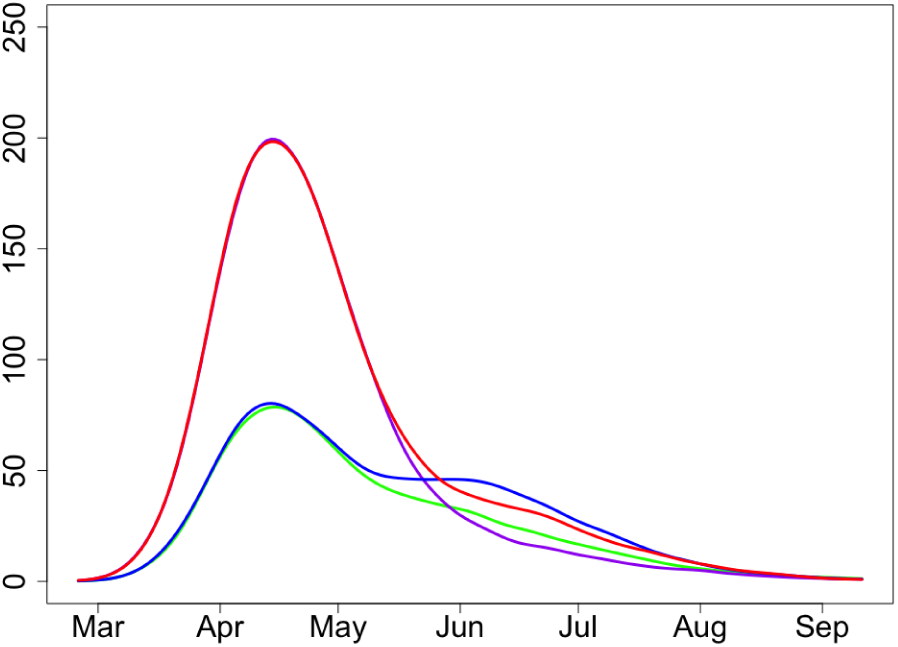
Expected deaths at time *t* at group *G* and the rest of population; *R*_2_ = 2, 2.5

Expected deaths toll for *R*_2_ = 2.5 is 16.9(5.9+11)K. This is significantly higher than 15(5.2+9.8)K for *R*_2_ = 2. This implies that the public should carry on some level of isolation in the next 2-3 months.

#### Conclusion

*Increase in the reproductive number after lifting the lock-down would inevitably imply a significant increase in the number of severe cases and expected death numbers*.

### 3.6 Sensitivity to *k*_*M*_ and *k*_*S*_, the shape parameter of the Erlang distributions for mild and severe cases

In Figures 22 and 23, we use *k*_*M*_ = 1, 3. The value *k*_*M*_ = 3 is default while the value *k*_*M*_ = 1 defines the exponential distribution for the period of infection in the case of mild disease and is equivalent to the corresponding assumption in SIR models. As the variance of the exponential distribution is larger than of the Erlang with *k*_*M*_ = 3 (given the same means), the epidemic with *k*_*M*_ = 1 runs longer and smoother.

**Figure 22:**
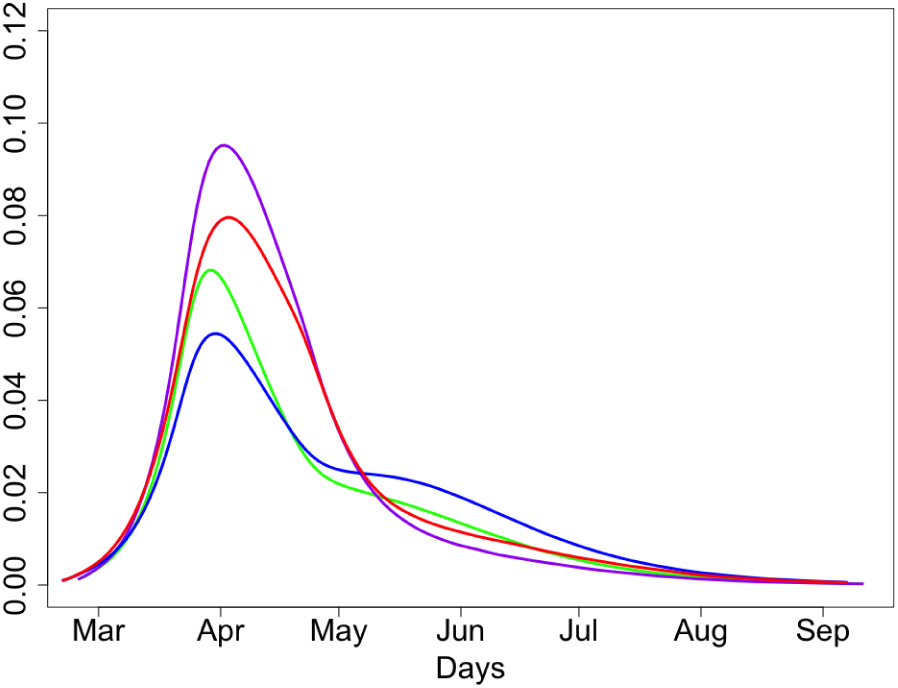
Proportions of people infected at time *t*; *k*_*M*_ = 1, 3

**Figure 23:**
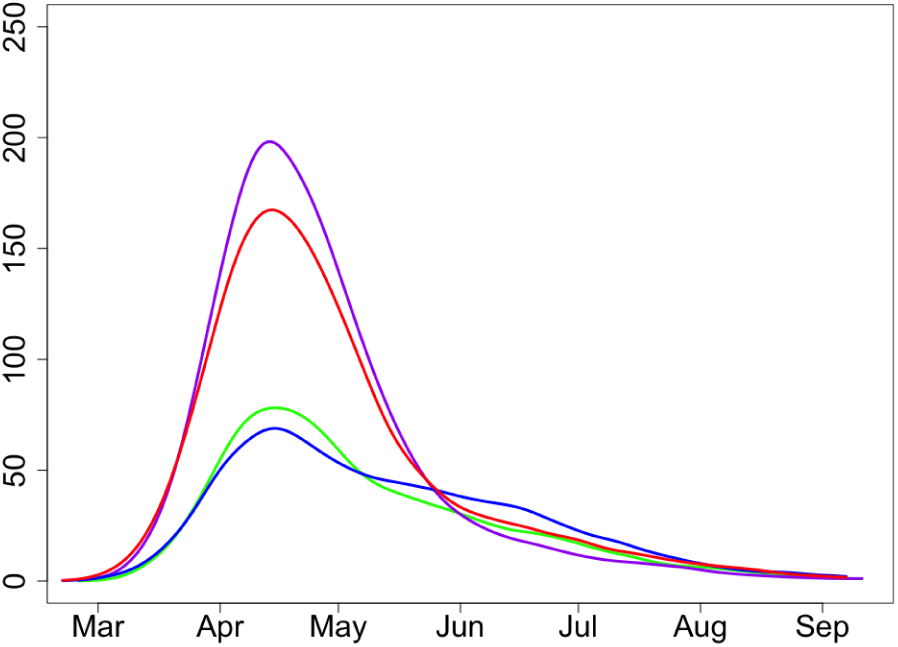
Expected deaths at time *t* at group *G* and the rest of population; *k*_*M*_ = 1, 3

In Figures 24 and 25, we use *k*_*S*_ = 1, 3. Parameter *k*_*S*_ is less sensitive than *k*_*M*_ (as there are less severe cases than the mild ones) but it is also is.

**Figure 24:**
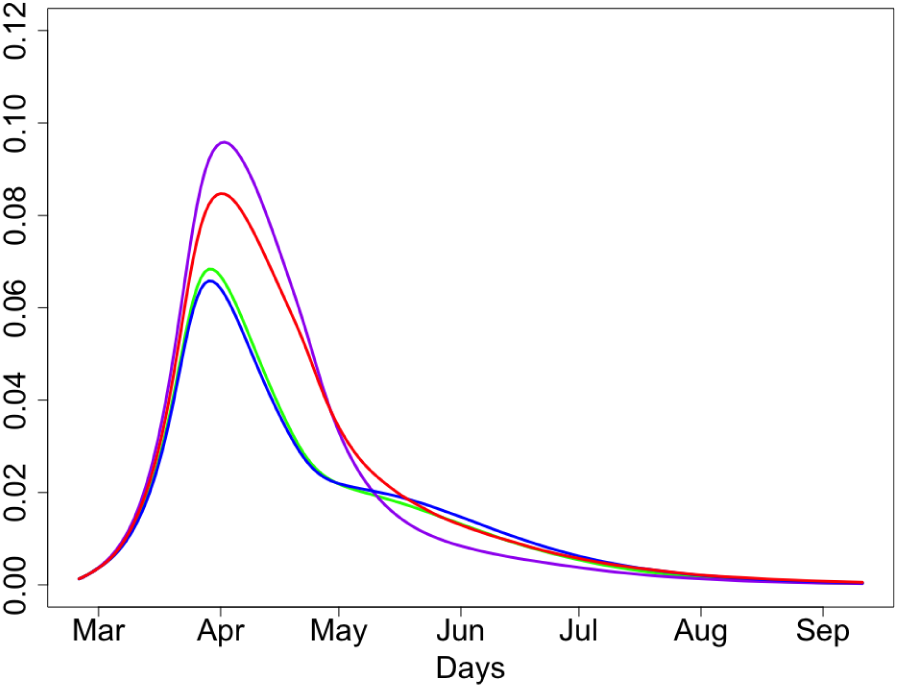
Proportions of people infected at time *t*; *k*_*S*_ = 1, 3

**Figure 25:**
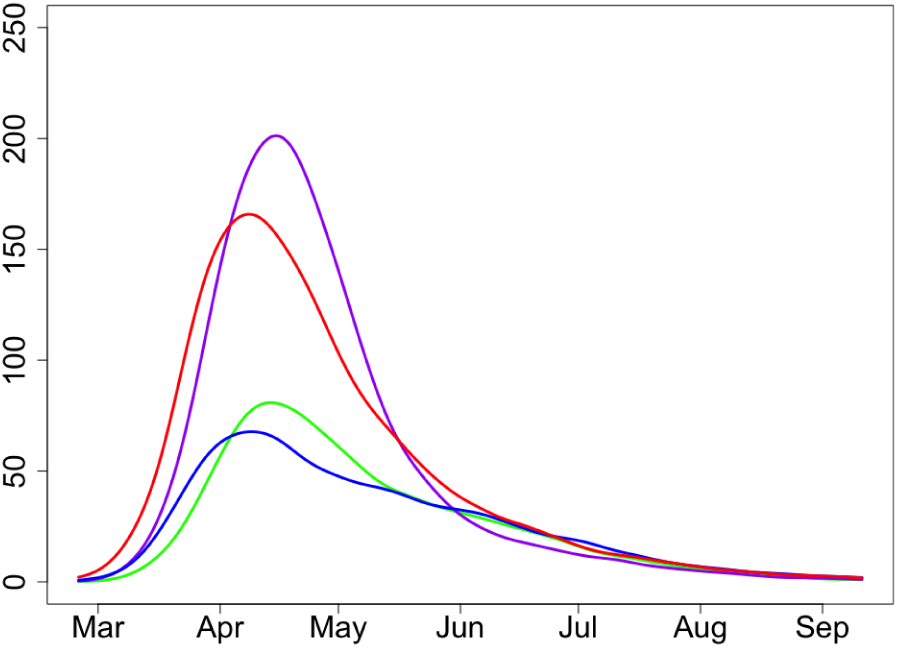
Expected deaths at time *t* at group *G* and the rest of population; *k*_*S*_ = 1, 3

#### Summary

*The parameter k*_*M*_ *is rather important and more information is needed about the distribution time of infectious period. Parameter k*_*S*_ *is less sensitive than k*_*M*_ *but it is also is*.

### 3.7 Sensitivity to *δ*, the probability of death in severe cases

In Figures 26 and 27, we use *δ* = 0.2 and 0.1. Decrease of *δ* increases the number of severe cases but does not change the expected death numbers.

**Figure 26:**
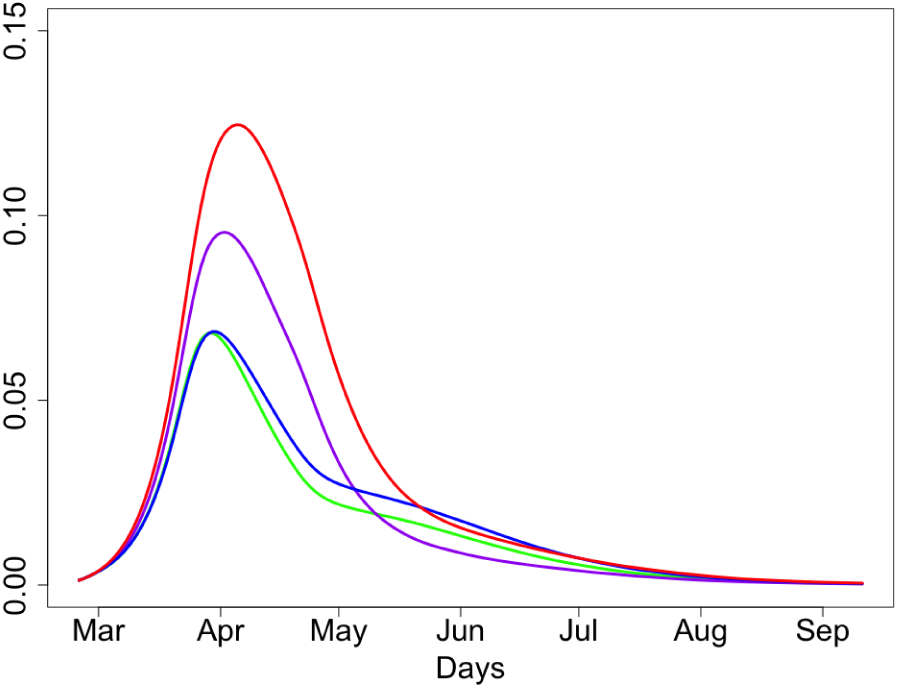
Proportions of people infected at time *t*; *δ* = 0.1, 0.2

**Figure 27:**
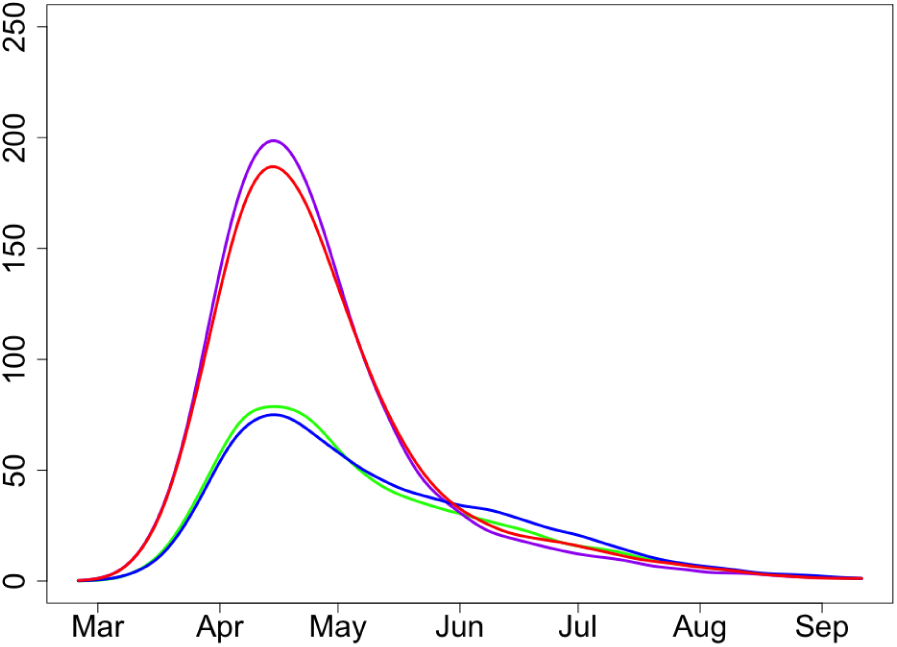
Expected deaths at time *t* at group *G* and the rest of population; *δ* = 0.1, 0.2

## Data Availability

The paper is based on simulation data only.

